# Exploring Physical and Biological Manifestations of Burnout and Post-Traumatic Stress Disorder Symptoms in Healthcare Workers: A Scoping Review Protocol

**DOI:** 10.1101/2023.04.16.23288657

**Authors:** Janey Kottler, Monica J. Gingell, Shaveta Khosla, Mitchell Kordzikowski, Rebecca Raszewski, David Chestek, Katherine A. Maki

## Abstract

**Introduction:** The COVID-19 pandemic has strained the mental and physical well-being of healthcare workers (HCW). Increased work-related stress and limited resources has increased symptoms of anxiety, depression, insomnia, and post-traumatic stress disorder (PTSD) in this population. Stress-related disorders have been strongly associated with long-term consequences including cardiometabolic disorders, endocrine disorders and premature mortality. This scoping review aims to explore available literature on burnout, PTSD, and other mental health-associated symptoms in HCW to synthesize relationships with physiological and biological biomarkers that may be associated with increased risk of disease, creating an opportunity to summarize current biomarker knowledge and identify gaps in this literature.

**Methods and Analysis:** This scoping review uses the Arksey and O’Malley six-step scoping review methodology framework. The research team will select appropriate primary sources using a search strategy developed in collaboration with a health sciences librarian. Three reviewers will initially screen the title and abstracts obtained from the literature searches, and two reviewers will conduct independent reviews of full-text studies for inclusion. The research team will be reviewing literature focusing on which burnout and/or PTSD-associated physiological and biological biomarkers have been studied, the methodologies used to study them and the correlations between the biomarkers and HCW experiencing burnout/PTSD. Data extraction forms will be completed by two reviewers for included studies and will guide literature synthesis and analysis to determine common themes.

**Ethics and Dissemination:** This review does not require ethical approval. We expect results from this scoping review to identify gaps in the literature and encourage future research regarding improving biologic and physiologic biomarker research in HCW. Preliminary results and general themes will be communicated back to stakeholders. Results will be disseminated through peer-reviewed publications, policy briefs, and conferences, as well as presented to stakeholders to an effort to invest in HCW mental and physical health.

**Strengths and Limitations of This Study:**

- This will be the first scoping review to assess the current understanding of the biologic and physiological impact of burnout on healthcare workers. The target population is restricted to healthcare workers; however, identified research gaps may be used to guide future studies in other high-burnout occupations and industries.
- This scoping review will be guided by the Arksey and O’Malley six-step methodological framework and the Preferred Reporting Items for Systematic Reviews and Meta-Analyses extension for Scoping Review checklist.
- Both peer reviewed manuscript and pre-prints/abstracts will be evaluated, but studies that have not been peer reviewed will be notated in the summary table. Conference abstracts are excluded.
- Preliminary and final themes and results identified by this scoping review will be communicated to stakeholders, including hospital staff and HCW, to ensure agreement with our interpretation and to convey knowledge gained with our population of interest.
- This review will advance the field’s current understanding of mechanisms connecting the burnout and pathogenic stress to biologic and physiologic outcomes in healthcare workers and provide researchers with gaps in the literature to inform opportunities for future research.

## INTRODUCTION

Burnout, by definition, is an occupational phenomenon characterized by emotional exhaustion, depersonalization, and a diminished sense of personal accomplishment (1). Though people in all occupations can experience burnout, burnout among healthcare workers (HCW) has been an ongoing, pervasive issue as a result of increasing work-related allostatic load and stress reactivity (2, 3). HCW burnout was considered to be at a “crisis level” well before the COVID-19 pandemic, with 35-54% of nurses and physicians, and 45-60% of medical students and residents reporting symptoms (2, 4). The COVID-19 pandemic further exacerbated HCW burnout in an already overburdened health system due to grueling workloads, psychological distress, and hazard exposure paired with limited resources and inadequate emotional support (2, 5-9). Burnout in the field of healthcare has become such a heated topic that the United States (US) Surgeon General issued an advisory to address the crisis (2).

On an individual level, HCW burnout has been associated with an increased incidence of anxiety, depression, insomnia, and post-traumatic stress disorder (PTSD) (2, 5-7). Though the current data on healthcare workers suicides is limited, it has been noted that physicians in the US are at a greater risk of suicide than physicians in other similar countries, especially when looking at female physicians compared to their male counterparts (10). Despite this higher risk of suicide, the likelihood of seeking help is low. Even before the COVID-19 pandemic, a survey performed on physicians showed that those with suicidal thoughts were less likely to seek mental health assistance than physicians without suicidal thoughts (11). In a field already predisposed to mental health issues, there needs to be a focus shift on preventing mental health issues and their effects (11).

Physical health is also noted to be impacted by burnout in HCW, with notable increases in chronic illnesses such as hypertension, diabetes mellitus and hyperlipidemia since the start of the COVID-19 pandemic (12). One study demonstrated that HCW experienced cardiac diseases and physical pain as aftermath from burnout (5). This may be explained by abnormal functioning and reactivity of both the autonomic nervous system (ANS) and hypothalamic-pituitary-adrenal (HPA) axis resulting in the cardiovascular system going into overdrive (e.g., heart rate and blood pressure); disrupting the body’s metabolic and immune systems(5). The cardiovascular system may also be impacted by other negative health implications of burnout including sleep dysfunction, endocrinologic disorders, and poor health behaviors and habits, along with direction associations between work stress and significant cardiovascular events (13, 14).

Although there are many hypotheses on how manifestations of work-associated burnout are associated with increased morbidity and mortality in HCW, until recently, the research on biobehavioral outcomes of burnout and symptoms of PTSD was limited as many studies solely emphasized the mental health implications of HCW burnout (15). On a systemic level, these health impacts on HCW are associated with poor patient outcomes and staffing shortages, leading to a cyclical impact of health care delivery systems; both posing significant risks to public health (2, 8, 9).

Though the mental impact of burnout on individuals is widely accepted, research studying associations between HCW burnout with the biologic and physiologic biomarkers that may inform mechanisms associated with increased risk of chronic disease remain inconclusive in current literature (16). In fact, biomarkers regarding mental and physical well-being have been understudied in many other high-stress professions at increased risk for occupational-related poor health outcomes as well, such as teachers, police officers, and lawyers. Broadly, a biomarker is a defined characteristic or objective measure that can be quantified accurately and reproducibly, and is an indicator of normal biologic and physiologic processes, pathogenic processes (as in disease) or responses to an exposure or intervention(13, 17-19).Biologic biomarkers are defined as a compound (e.g., molecule, hormone, metabolite, microbe) that is found in the blood, body fluids or tissues that is an objective measure of homeostatic or pathologic functioning of an organism (4, 20)(see **Table 1**). In research, biologic biomarkers are usually collected in blood samples, saliva samples and urine samples, but the incorporation of microbiome- and metabolite-based biologic biomarkers in biobehavioral research has expanded samples collected to alternative tissues like fecal samples, skin swabs and dental plaque samples (21, 22). Physiologic biomarkers have not been as explicitly defined in the literature but are objective markers or indicators of the functioning and status of the human body, often stemming from measures of sleep or vital signs (**Table 1**). Sleep and activity-based physiologic biomarkers are usually measured with actigraphy or consumer-driven products (e.g., Fitbit, WHOOP). Actigraphy records and integrates limb movement activity by quantifying acceleration and devices are designed to be worn on the waist, wrist or ankle (23). Conversely, in consumer-based sleep and activity recording devices, data can be collected from a range of methods including actigraphy, geographic location, skin conductivity, and noise levels, showing a lack of standardization. Consumer-based devices also have a wide range of platforms used for data collection including devices classified as wearables worn directly by the user (e.g., watch-based devices, rings) or nearables where the device is proximal but not in direct contact with the user (phone-based collection devices or other platforms). Physiologic biomarkers can also be collected by ambulatory monitoring systems (e.g. home blood pressure monitors) or electrocardiogram monitoring. These wearable devices may assist in obtaining the foundation for burnout biomarkers, increasing the quality of individualized diagnostic and treatment plans for HCW experiencing burnout (14). Understanding the underlying biomarkers impacting HCW can provide significant advances to the treatment of burnout, potentially improving quality of life (16).

**Table 1.**
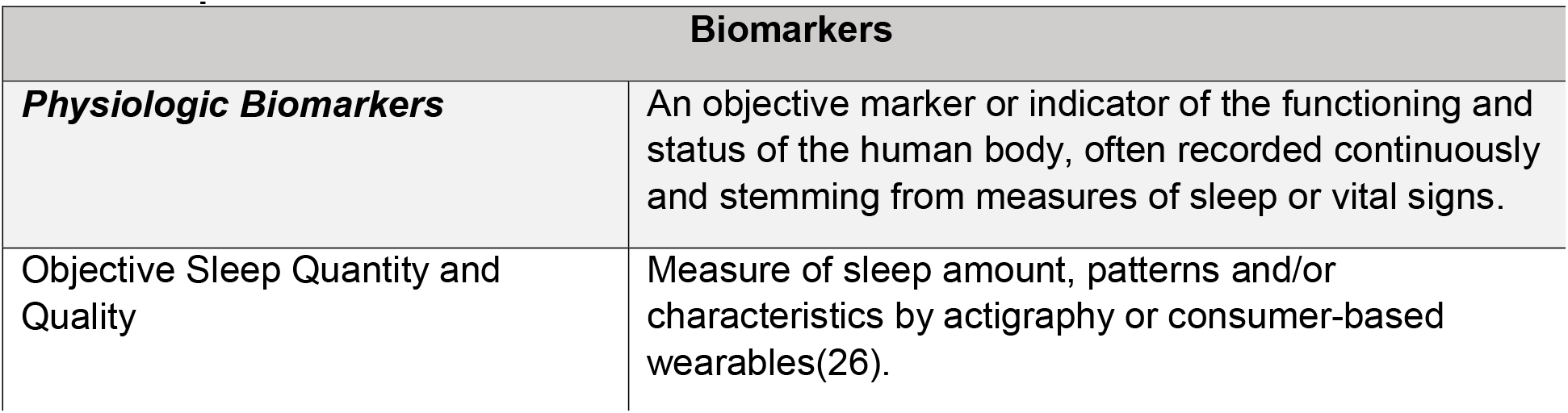
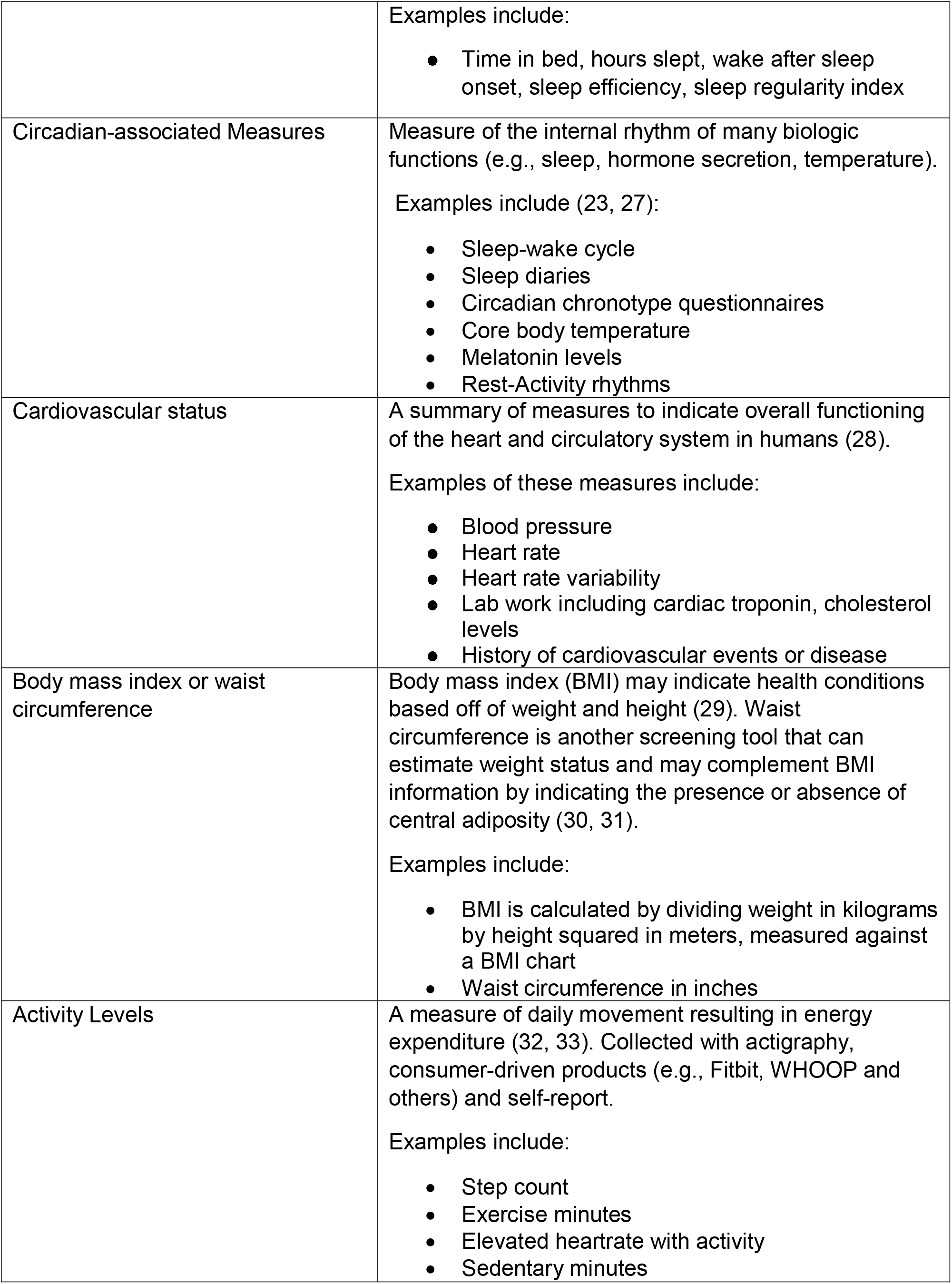
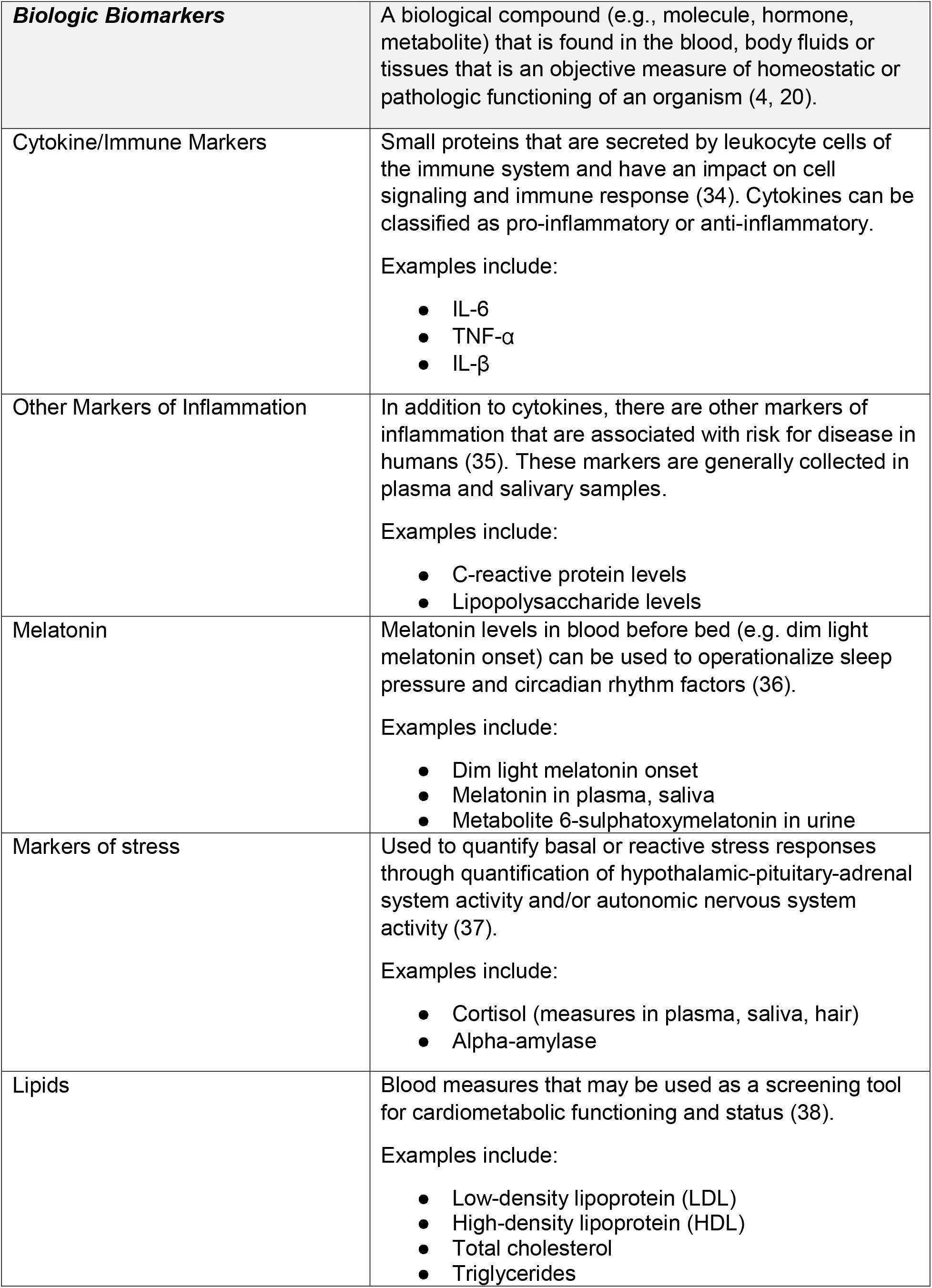
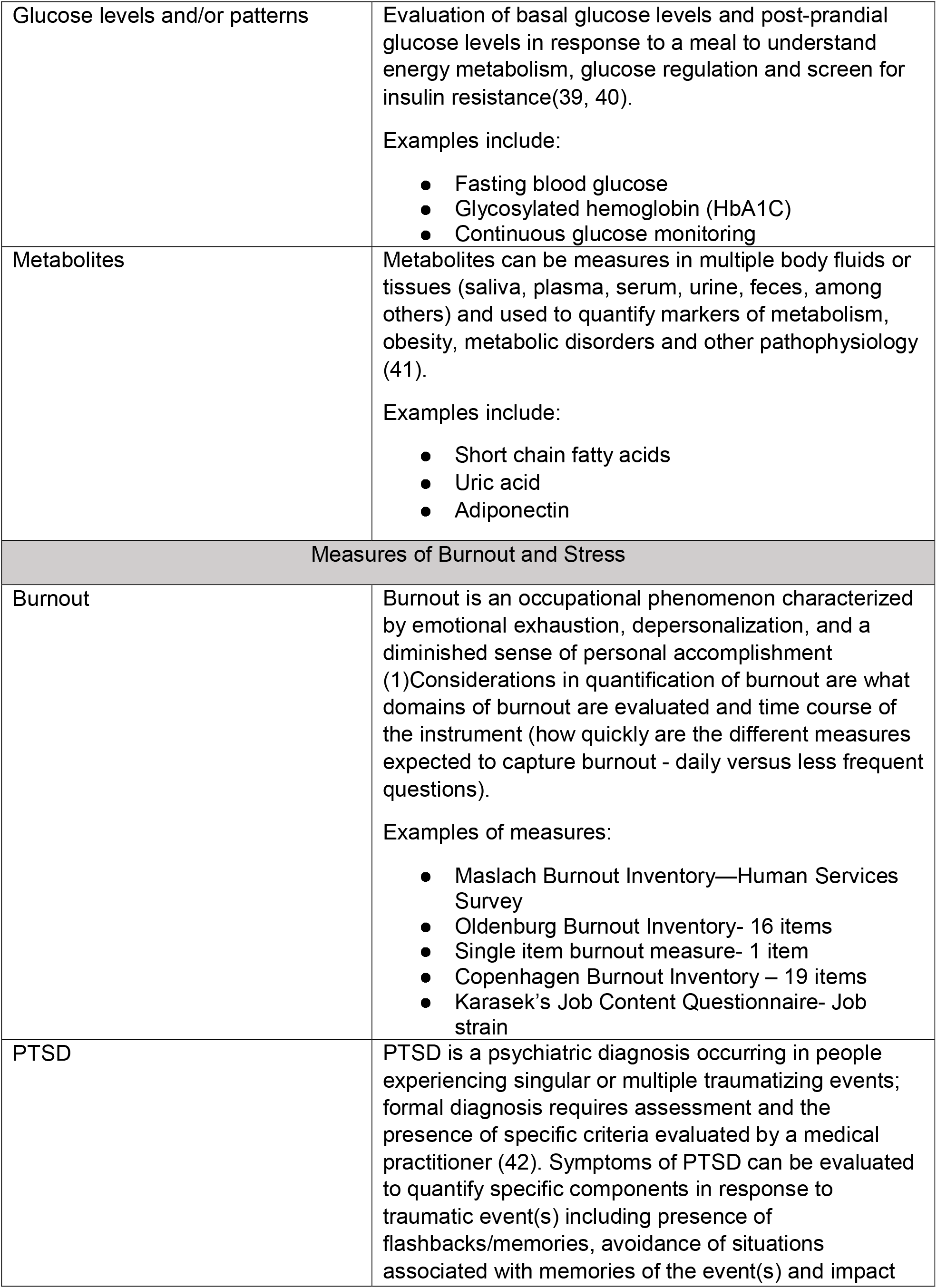
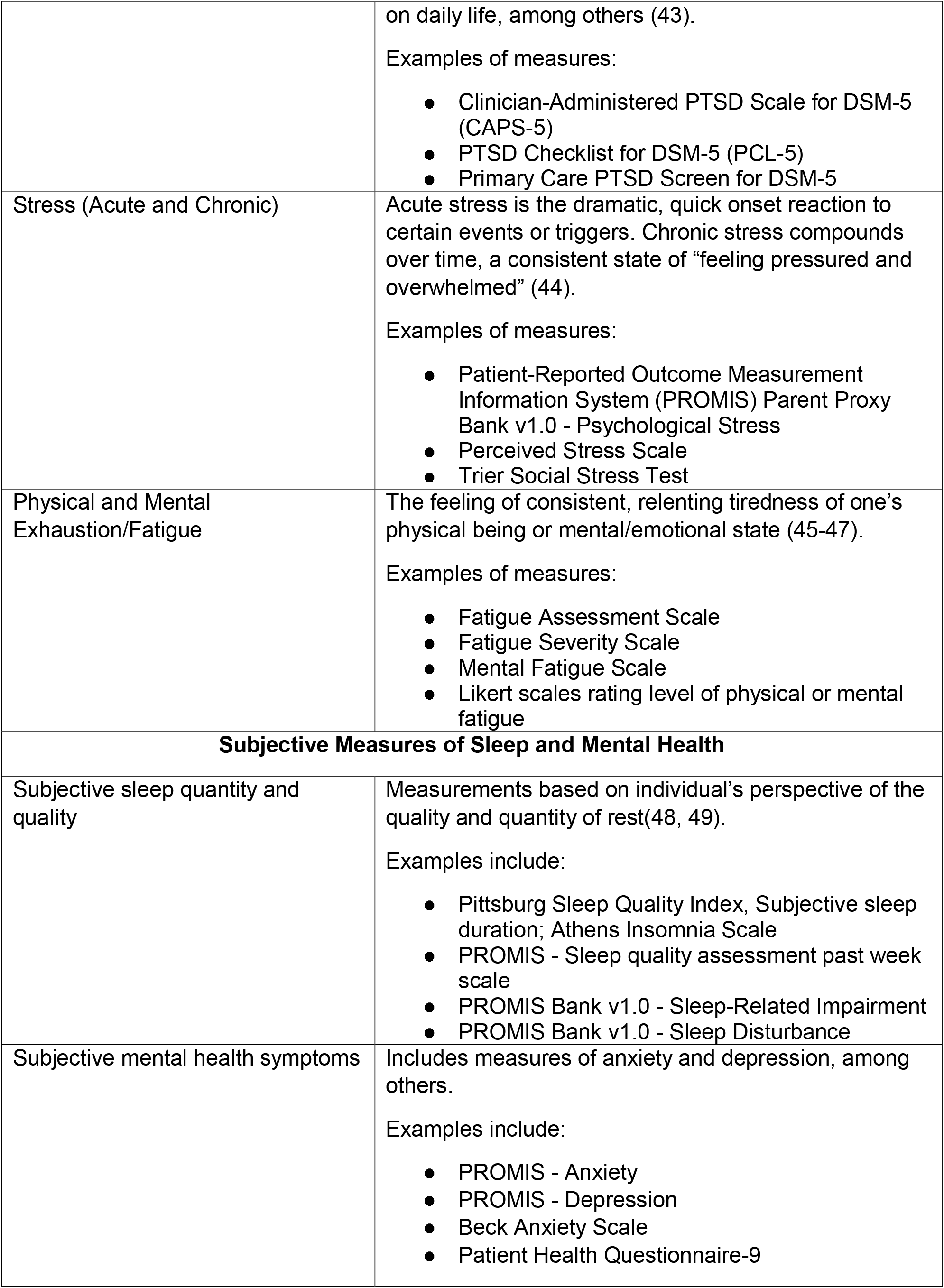
Operational Definitions.

| **Biomarkers** |  |
| --- | --- |
| ***Physiologic Biomarkers*** | An objective marker or indicator of the functioning and status of the human body, often recorded continuously and stemming from measures of sleep or vital signs. |
| Objective Sleep Quantity and Quality | Measure of sleep amount, patterns and/or characteristics by actigraphy or consumer-based wearables(26). |
|  | Examples include:<br>• Time in bed, hours slept, wake after sleep onset, sleep efficiency, sleep regularity index |
| Circadian-associated Measures | Measure of the internal rhythm of many biologic functions (e.g., sleep, hormone secretion, temperature).<br>Examples include (23, 27):<br>• Sleep-wake cycle<br>• Sleep diaries<br>• Circadian chronotype questionnaires<br>• Core body temperature<br>• Melatonin levels<br>• Rest-Activity rhythms |
| Cardiovascular status | A summary of measures to indicate overall functioning of the heart and circulatory system in humans (28).<br>Examples of these measures include:<br>• Blood pressure<br>• Heart rate<br>• Heart rate variability<br>• Lab work including cardiac troponin, cholesterol levels<br>• History of cardiovascular events or disease |
| Body mass index or waist circumference | Body mass index (BMI) may indicate health conditions based off of weight and height (29). Waist circumference is another screening tool that can estimate weight status and may complement BMI information by indicating the presence or absence of central adiposity (30, 31).<br>Examples include:<br>• BMI is calculated by dividing weight in kilograms by height squared in meters, measured against a BMI chart<br>• Waist circumference in inches |
| Activity Levels | A measure of daily movement resulting in energy expenditure (32, 33). Collected with actigraphy, consumer-driven products (e.g., Fitbit, WHOOP and others) and self-report.<br>Examples include:<br>• Step count<br>• Exercise minutes<br>• Elevated heartrate with activity<br>• Sedentary minutes |
| **Biologic Biomarkers** | A biological compound (e.g., molecule, hormone, metabolite) that is found in the blood, body fluids or tissues that is an objective measure of homeostatic or pathologic functioning of an organism (4, 20). |
| Cytokine/Immune Markers | Small proteins that are secreted by leukocyte cells of the immune system and have an impact on cell signaling and immune response (34). Cytokines can be classified as pro-inflammatory or anti-inflammatory.<br>Examples include:<br>• IL-6<br>• TNF-α<br>• IL-β |
| Other Markers of Inflammation | In addition to cytokines, there are other markers of inflammation that are associated with risk for disease in humans (35). These markers are generally collected in plasma and salivary samples.<br>Examples include:<br>• C-reactive protein levels<br>• Lipopolysaccharide levels |
| Melatonin | Melatonin levels in blood before bed (e.g. dim light melatonin onset) can be used to operationalize sleep pressure and circadian rhythm factors (36).<br>Examples include:<br>• Dim light melatonin onset<br>• Melatonin in plasma, saliva<br>• Metabolite 6-sulphatoxymelatonin in urine |
| Markers of stress | Used to quantify basal or reactive stress responses through quantification of hypothalamic-pituitary-adrenal system activity and/or autonomic nervous system activity (37).<br>Examples include:<br>• Cortisol (measures in plasma, saliva, hair)<br>• Alpha-amylase |
| Lipids | Blood measures that may be used as a screening tool for cardiometabolic functioning and status (38).<br>Examples include:<br>• Low-density lipoprotein (LDL)<br>• High-density lipoprotein (HDL)<br>• Total cholesterol<br>• Triglycerides |
| Glucose levels and/or patterns | Evaluation of basal glucose levels and post-prandial glucose levels in response to a meal to understand energy metabolism, glucose regulation and screen for insulin resistance(39, 40).<br>Examples include:<br>• Fasting blood glucose<br>• Glycosylated hemoglobin (HbA1C)<br>• Continuous glucose monitoring |
| Metabolites | Metabolites can be measures in multiple body fluids or tissues (saliva, plasma, serum, urine, feces, among others) and used to quantify markers of metabolism, obesity, metabolic disorders and other pathophysiology (41).<br>Examples include:<br>• Short chain fatty acids<br>• Uric acid<br>• Adiponectin |
| Measures of Burnout and Stress |  |
| Burnout | Burnout is an occupational phenomenon characterized by emotional exhaustion, depersonalization, and a diminished sense of personal accomplishment (1)Considerations in quantification of burnout are what domains of burnout are evaluated and time course of the instrument (how quickly are the different measures expected to capture burnout - daily versus less frequent questions).<br>Examples of measures:<br>• Maslach Burnout Inventory—Human Services Survey<br>• Oldenburg Burnout Inventory- 16 items<br>• Single item burnout measure- 1 item<br>• Copenhagen Burnout Inventory – 19 items<br>• Karasek's Job Content Questionnaire- Job strain |
| PTSD | PTSD is a psychiatric diagnosis occurring in people experiencing singular or multiple traumatizing events; formal diagnosis requires assessment and the presence of specific criteria evaluated by a medical practitioner (42). Symptoms of PTSD can be evaluated to quantify specific components in response to traumatic event(s) including presence of flashbacks/memories, avoidance of situations associated with memories of the event(s) and impact |
|  | on daily life, among others (43).<br>Examples of measures:<br>• Clinician-Administered PTSD Scale for DSM-5 (CAPS-5)<br>• PTSD Checklist for DSM-5 (PCL-5)<br>• Primary Care PTSD Screen for DSM-5 |
| Stress (Acute and Chronic) | Acute stress is the dramatic, quick onset reaction to certain events or triggers. Chronic stress compounds over time, a consistent state of “feeling pressured and overwhelmed” (44).<br>Examples of measures:<br>• Patient-Reported Outcome Measurement Information System (PROMIS) Parent Proxy Bank v1.0 - Psychological Stress<br>• Perceived Stress Scale<br>• Trier Social Stress Test |
| Physical and Mental Exhaustion/Fatigue | The feeling of consistent, relenting tiredness of one’s physical being or mental/emotional state (45-47).<br>Examples of measures:<br>• Fatigue Assessment Scale<br>• Fatigue Severity Scale<br>• Mental Fatigue Scale<br>• Likert scales rating level of physical or mental fatigue |
| **Subjective Measures of Sleep and Mental Health** |  |
| Subjective sleep quantity and quality | Measurements based on individual’s perspective of the quality and quantity of rest(48, 49).<br>Examples include:<br>• Pittsburg Sleep Quality Index, Subjective sleep duration; Athens Insomnia Scale<br>• PROMIS - Sleep quality assessment past week scale<br>• PROMIS Bank v1.0 - Sleep-Related Impairment<br>• PROMIS Bank v1.0 - Sleep Disturbance |
| Subjective mental health symptoms | Includes measures of anxiety and depression, among others.<br>Examples include:<br>• PROMIS - Anxiety<br>• PROMIS - Depression<br>• Beck Anxiety Scale<br>• Patient Health Questionnaire-9 |

Due to the only recently emerging literature studying the biologic and physiologic associations with burnout and PTSD in HCW, along with no known synthesis of these biomarker responses with symptoms of burnout and PTSD in HCW, a scoping review is deemed necessary. Furthermore, the quantification of burnout in HCW is also based on a number of subjective instruments focusing on different domains of stress and burnout, which also poses a need to survey the current literature on how the concept of subjective burnout and PTSD is being evaluated in HCW(24, 25) to quantify current practices and evaluate gaps, if present. Therefore, the aims of this scoping review include: 1) understanding the research methodologies utilized in the literature when studying biologic and physiologic biomarkers associations with burnout and/or PTSD in HCW; 2) assessing which of these biomarkers have been studied in HCW experiencing burnout and/or PTSD; and 3) determining reported associations between burnout and/or PTSD and these biomarkers in HCW.

## METHODS/DESIGN

### protocol design

This scoping review will use a methodology framework developed by Arksey and O’Malley and enhanced by Levac, Colquhoun, and O’Brien (50, 51). This step-by-step methodology consists of five stages: 1) identifying the research questions, 2) identifying relevant studies, 3) selecting studies, 4) charting the data, and 5) collating, summarizing and reporting the results. The Preferred Reporting Items for Systematic Reviews and Meta-Analyses extension for Scoping Review (PRISMA-ScR) checklist will be used throughout the review to guide our search strategy, article selection, and result synthesis (52).

#### Stage 1: Identifying the research question

The overarching aim of this review is to understand known associations between biologic and physiologic biomarkers and burnout and PTSD in HCW, as well as highlight corresponding gaps in the literature. The research team defined the following three research questions:

- What methodologies have been used to quantify subjective burnout/PTSD and study relationships between burnout/PTSD and biologic and physiologic biomarkers in HCW?
- What biologic and physiologic biomarkers have been studied in HCW experiencing burnout/PTSD?
- What biologic and physiologic biomarkers are positively or negatively associated with burnout/PTSD in HCW?

#### Stage 2: Identifying relevant studies

Initial search strategies for this review were developed in collaboration with a health sciences librarian to develop comprehensive terms constructed to capture our variables of interest (e.g. burnout, symptoms of PTSD, biological biomarkers, and physiological biomarkers). When the search strategy is finalized, subject and keyword searches will be conducted within the following databases: APA PsycINFO via ProQuest, CINAHL Plus with Full Text, Embase: Excerpta Medica Database via Elsevier, and Medline via Ovid. Initial subject headings and keyword searches were built for the following concepts: 1. health care professionals who worked in inpatient and outpatient settings; 2. psychological and physiological stress, traumatic stress disorders and fatigue; and 3. biological and physiological biomarkers. Search filters on health care professionals and students built by a librarian at the University of Alberta were adapted and customized for each database (53).

The current limitations include: target population restricted to HCW including nurses, physicians, allied health professionals and other inpatient/outpatient hospital staff. After the subject headings and keyword searches for each concept are combined together, the search results will be limited to English and the years 2013-2023 within all the databases. We are focusing on literature completed over the last 10 years as much of the technology to quantify biologic and physiology biomarkers rapidly evolves with time and we want to focus synthesis results and recommendations on biomarkers that will be relevant in future research. Dissertations and conference abstracts will be excluded from CINAHL and Embase, respectively. In APA PsycINFO, search results will be limited to Scholarly Journals to automatically exclude books, book chapters, and dissertations from the search. Once final searches are completed, the citations will be uploaded into the screening tool, Covidence (Covidence systematic review software, Veritas Health Innovation, Melbourne, Australia. Available at www.covidence.org), in order to remove duplicates and initiate citation screening.

#### Stage 3: Selecting studies

This scoping review will consider study designs including both experimental and observational study designs, although we anticipate a majority of research focused on relationships between HCW burnout/PTSD and biologic and physiologic biomarkers will have an observational study design. Nevertheless, the included study designs will be randomized controlled trials, non-randomized controlled trials, and meta-analyses, along with analytical and descriptive observational studies including cohort studies, case-control studies, cross-sectional studies, case series, and case reports. Qualitative/mixed methods studies will be included if the results also include comparisons between burnout/symptoms of PTSD and physiologic or biologic biomarkers.

Covidence software will be used to store and organize all articles being reviewed, including initial data screening, selection and data extraction. Initially, three research team members will initially pilot-test the search strategy, screening the title and abstract on a small number of citations (e.g., about six to eight citations), and make search strategy adjustments based on reviewer feedback. Thereafter, screening will continue for the resulting articles. If more than 5 conflicts between screening team members are identified, title and abstract screening will stop and the research team will meet to discuss screening criteria so agreement is met and inter-rater reliability is increased. For articles passing title/abstract screening, two research team members will review the included full-text articles, deciding which meet the inclusion criteria. Disagreements that arise between the team members will be resolved through discussion and a third research team member via independent review. If full text articles are excluded after review, we will document the reason for exclusion in line with PRISMA-ScR guidelines. Reviewing all articles by the research term using the protocol outlined in this manuscript will take approximately three-four months to complete.

Article inclusion criteria will include the population of human research participants older than 18 years of age; occupation of HCW; focusing on HCW burnout, PTSD or both; published between 2013 to 2023; outcome measures related to biomarker associations (see **Table 1**), and articles published in English (irrespective of the geographic location of the study). The research team will consider articles including other mental health symptoms if they also include HCW burnout and PTSD focus. We will also include articles that look at interventions utilized for HCW burnout or PTSD if biologic and physiologic biomarkers are the outcomes. Articles excluded from this scoping review would include those not available in English; review articles; conference abstracts and presentations; and outcome measures not related to the variables outlined in **Table 1**.

#### Stage 4: Charting the data

A standardized table of included studies and associated source data will be recorded throughout the synthesis process after studies are selected. The data extraction form template was created by the research team to synthesize and highlight different aspects of each article. Two study team members will pilot the data charting form for the first 5 articles that meet full text inclusion criteria and will update it as necessary after discussion with the research team. See **Table 2** for the currently proposed data extraction form template. Charting the pertinent primary source manuscript components utilizing the form in **Table 2** will take approximately three to six months depending on the number of full text articles that meet criteria for data extraction.

**Table 2.**
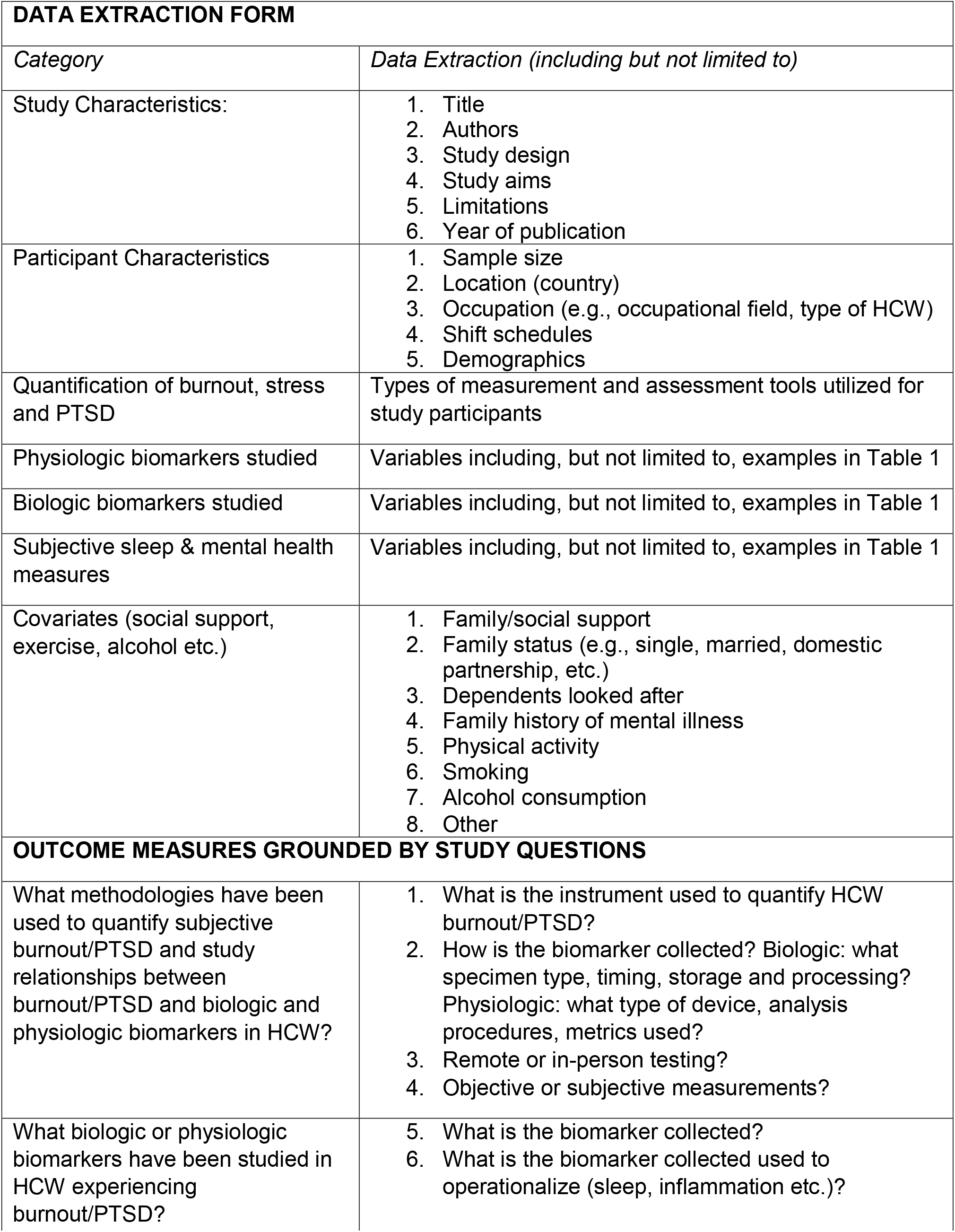
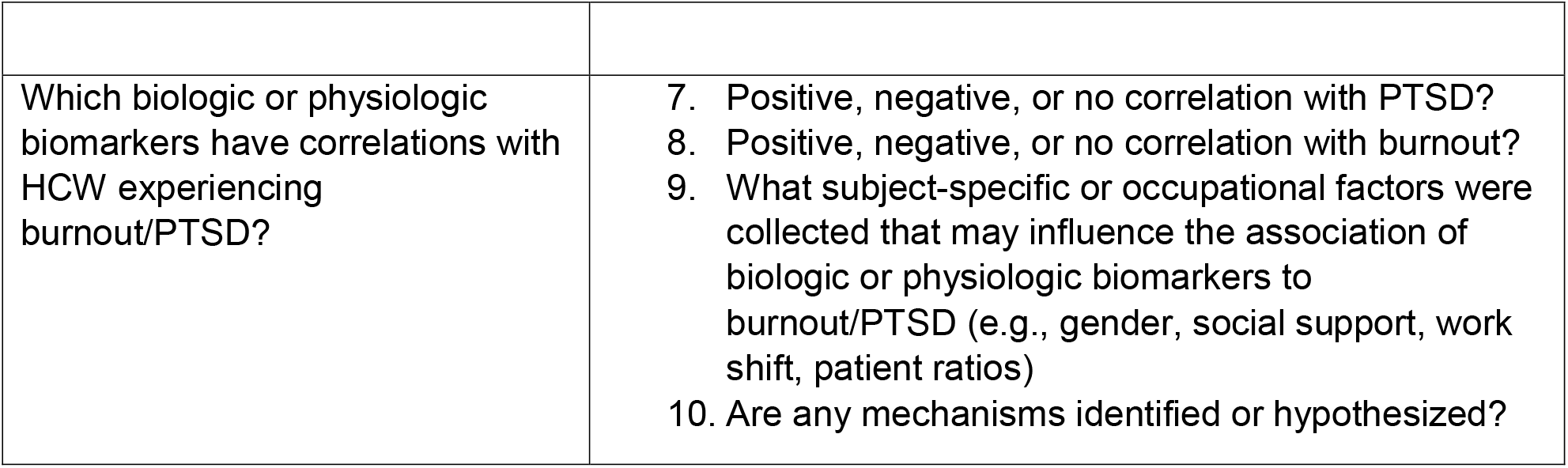
Data Charting Form.

| **DATA EXTRACTION FORM** |  |
| --- | --- |
| *Category* | *Data Extraction (including but not limited to)* |
| Study Characteristics: | 1. Title<br>2. Authors<br>3. Study design<br>4. Study aims<br>5. Limitations<br>6. Year of publication |
| Participant Characteristics | 1. Sample size<br>2. Location (country)<br>3. Occupation (e.g., occupational field, type of HCW)<br>4. Shift schedules<br>5. Demographics |
| Quantification of burnout, stress and PTSD | Types of measurement and assessment tools utilized for study participants |
| Physiologic biomarkers studied | Variables including, but not limited to, examples in Table 1 |
| Biologic biomarkers studied | Variables including, but not limited to, examples in Table 1 |
| Subjective sleep & mental health measures | Variables including, but not limited to, examples in Table 1 |
| Covariates (social support, exercise, alcohol etc.) | 1. Family/social support<br>2. Family status (e.g., single, married, domestic partnership, etc.)<br>3. Dependents looked after<br>4. Family history of mental illness<br>5. Physical activity<br>6. Smoking<br>7. Alcohol consumption<br>8. Other |
| **OUTCOME MEASURES GROUNDED BY STUDY QUESTIONS** |  |
| What methodologies have been used to quantify subjective burnout/PTSD and study relationships between burnout/PTSD and biologic and physiologic biomarkers in HCW? | 1. What is the instrument used to quantify HCW burnout/PTSD?<br>2. How is the biomarker collected? Biologic: what specimen type, timing, storage and processing? Physiologic: what type of device, analysis procedures, metrics used?<br>3. Remote or in-person testing?<br>4. Objective or subjective measurements? |
| What biologic or physiologic biomarkers have been studied in HCW experiencing burnout/PTSD? | 5. What is the biomarker collected?<br>6. What is the biomarker collected used to operationalize (sleep, inflammation etc.)? |
| Which biologic or physiologic biomarkers have correlations with HCW experiencing burnout/PTSD? | 7. Positive, negative, or no correlation with PTSD?<br>8. Positive, negative, or no correlation with burnout?<br>9. What subject-specific or occupational factors were collected that may influence the association of biologic or physiologic biomarkers to burnout/PTSD (e.g., gender, social support, work shift, patient ratios)<br>10. Are any mechanisms identified or hypothesized? |

In addition to the biologic and physiologic measures defined and outlined in **Table 1**, the data extraction forms will also compile demographic data, HCW shift schedules, measures of emotional support, smoking status and alcohol/drug use patterns.

#### Stage 5: Collating, summarizing and reporting the results

Results will be summarized with both qualitative and quantitative analyses, with the opportunity to display data in both tables or graphs. The results will be sub-analyzed according to which research questions were addressed. While challenging to compare and contrast biomarker outcomes in different HCW, the study team will highlight the specific HCW population (e.g., doctors, nurses) being studied, the type of biomarker assessed, as well as the specific study population’s occupational setting (e.g., inpatient, outpatient). **Table 2** highlights the specific categories our study team will extract from each full-text article, if they are reported.

#### Stage 6: Consultation

Though an optional step in Arksey and O’Malley’s scoping review framework, there is significance in directly involving stakeholders in discussions and implications of this review’s results. Stakeholders may include HCW, hospital administration, experts in this HCW mental and physical health, employee/occupational health teams and those with invested interest in HCW. Through personal relationships throughout the hospital system, results will be shared and disseminated in an effort to bring recognition to the mental and physical toll of our HCW. The goal would be to create agreements and interpretations of the scoping review’s results amongst stakeholders and create a feasible and sustainable action plan for protecting and investing in our HCW in the future.

## ETHICS AND DISSEMINATION

The summarized results from this scoping review will provide relevant stakeholders with a deeper understanding of the biologic and physiologic impact of burnout and PTSD on HCW and lead to future research developing systems-level interventions to mitigate HCW burnout and PTSD. This review does not involve access to individual-level data; therefore, it does not require ethical approval.

We plan to disseminate the results through peer-reviewed publications, policy briefs, and presentations at conferences and to stakeholders. It will be the responsibility of the research team to communicate findings to local healthcare institutions to draw awareness to the biomarkers impacted by mental health disorders in HCW.

### Patient and Public Involvement

The research team will not need patient or public involvement for the proposed scoping review.

## CONCLUSION

Our protocol for a scoping review to synthesize the current literature focused on associations between burnout and symptoms of PTSD with physiological and biological biomarkers in HCW roles is presented. The scoping review will comprehensively analyze existing literature for correlations between biomarkers and burnout and/or PTSD in HCW, assess methodologies used in biomarker research, and identify gaps in the literature to guide future research.

Intervening before or when HCW begin experiencing symptoms of burnout, trauma and PTSD can hopefully reduce occupational-associated HCW chronic disease, shortage, mental illness, and suicide. Therefore, a critical next step in addressing this issue is synthesizing the currently available literature to understand the state of the current science and stimulate future hypotheses regarding improving biomarker research in HCW. Understanding comprehensively how HCW are impacted biologically and physiologically by burnout, PTSD and potentially other mental health disorders can begin the conversation for how to protect HCW and intervene at appropriate times.

## Supporting information

Supplemental Table: Search strategy

Publishing support

## Data Availability

All data produced in the present work are contained in the manuscript.

## Abbreviations

BMI: Body mass index
COVID-19: Coronavirus Disease 2019
HCW: Healthcare Workers
PTSD: Post-Traumatic Stress Disorder
PRISMA-ScR: Preferred Reporting Items for Systematic Reviews and Meta-Analyses extension for Scoping Review
PROMIS: Patient-Reported Outcomes Measurement Information System
US: United States

## ACKNOWLEDGEMENTS

AUTHOR’S CONTRIBUTIONS: Conception or design of the work: JK, MG, KM Drafting and editing of manuscript: JK, MK, MG, SK, KM; Critical editing: RR, DC; Final approval: JK, MK, MG, RR, SK, DC, KM; Agreement to be accountable for all aspects of the work: JK, MK, MG, RR, SK, DC, KM. Sandy M. Campbell is also acknowledged for her original work with the health sciences professionals and students search filters.

