## Supplemental Table: Search strategy for "Exploring Physical and Biological Manifestations of Burnout and Post-Traumatic Stress Disorder Symptoms in Healthcare Workers: A Scoping Review Protocol"

### **Supplemental Table 1: Database-Specific Search Strategy**

**Database: APA PsycINFO
Provider: ProQuest**

Limits: English, Scholarly Journals, Years 2013-2023

|  | Concept | Search Strategy |
| --- | --- | --- |
| #1 | Health Care Professionals | ((((AB,TI,IF("allied health profession*" OR "allied health personnel*" OR "community health worker*" OR counsellor* OR counselor* OR "emergency medical technician*" OR "first responder" OR ("health educator" OR "health educators") OR (("healthcare assistant")) OR "healthcare support worker*" OR "home health aide*" OR "licensed practical nurse*" OR LPN OR nurse-aide* OR ("occupational therapist" OR "occupational therapists") OR paramedic* OR "paediatric assistant*" OR "pediatric assistant*" OR ("pharmacy technician" OR "pharmacy technicians") OR ("physical therapist" OR "physical therapists") OR "physical therapy assistant*" OR ("physician assistant" OR "physician assistants") OR "psychiatric aide*" OR ("respiratory therapist" OR "respiratory therapists") OR ("speech pathologist" OR "speech pathologists")) OR ((MAINSUBJECT.EXACT("Medical Personnel") OR MAINSUBJECT.EXACT.EXPLODE("Psychiatric Hospital Staff") OR MAINSUBJECT.EXACT("Clinicians") OR MAINSUBJECT.EXACT.EXPLODE("Allied Health Personnel") OR MAINSUBJECT.EXACT.EXPLODE("Nurses") OR MAINSUBJECT.EXACT("Clinical Psychologists") OR MAINSUBJECT.EXACT.EXPLODE("Therapists") OR MAINSUBJECT.EXACT("Mental Health Personnel") OR MAINSUBJECT.EXACT("Optometrists") OR MAINSUBJECT.EXACT.EXPLODE("Psychotherapists") OR MAINSUBJECT.EXACT("Health Personnel") OR MAINSUBJECT.EXACT.EXPLODE("Physicians") OR MAINSUBJECT.EXACT("Psychologists") OR MAINSUBJECT.EXACT("Pharmacists")) OR MAINSUBJECT.EXACT("Paramedics") OR MAINSUBJECT.EXACT.EXPLODE("counselors") OR MAINSUBJECT.EXACT.EXPLODE("social workers") OR AB,TI,IF(allergist* OR anesthesiologist* OR anesthetist* OR attending* OR cardiologist* OR clinician* OR dermatologist* OR diabetologist* OR doctor* OR emergency physician* OR endocrinologist* OR fellow OR fellows OR gastroenterologist* OR ("general practitioner" OR "general practitioners") OR geriatrician* OR gynecologist* OR hematologist* OR haematologist* OR hospitalist* OR internist* OR ("medical resident" OR "medical residents") OR neonatologist* OR nephrologist* OR neurologist* OR neurosurgeon* OR nurse OR nurses OR obstetrician* OR oncologist* OR ophthalmologist* OR oral surgeon* OR osteopath OR osteopaths OR otolaryngologist* OR pediatrician* OR pharmacist* OR phlebotomist* OR physician* OR podiatrist* OR psychologist* OR psychiatrist* OR pulmonologist* OR rheumatologist* OR ("social worker" OR "social workers") OR surgeon* OR therapist* OR toxicologist* OR urologist* OR immunologist* OR hepatologist* OR internist* OR intensivist* OR gynaecologist* OR paediatrician*)))) OR (AB,TI,IF((health OR "health care" OR healthcare OR medical) AND (worker* OR practitioner* OR provider* OR professional* OR navigator* OR staff* OR personnel*))))) |
| #2 | Psychological and Physiological Stress, Traumatic Stress Disorders, & Fatigue | (((MAINSUBJECT.EXACT("Caregiver Burden") OR MAINSUBJECT.EXACT("Acute Stress Disorder") OR ((MAINSUBJECT.EXACT("Fatigue")) OR MAINSUBJECT.EXACT("Posttraumatic Stress") OR MAINSUBJECT.EXACT("Psychological Stress") OR MAINSUBJECT.EXACT("Occupational Stress") OR MAINSUBJECT.EXACT.EXPLODE("Physiological Stress")) OR (MAINSUBJECT.EXACT.EXPLODE("Posttraumatic Stress Disorder") OR MAINSUBJECT.EXACT("Stress and Trauma Related Disorders"))) OR AB,TI,IF("mental stress" OR "psychological trauma*" OR PTSD OR "burn out*" OR burnout* OR "occupational stress*" OR "job stress*" OR "workplace stress*" OR exhaustion) OR (TITLE-ABS (trauma* W/3 disorder)))) |
| #3 | Physiological Biomarkers | ((((MAINSUBJECT.EXACT.EXPLODE("Sleep") OR MAINSUBJECT.EXACT("Sleep Deprivation") OR MAINSUBJECT.EXACT("Sleepiness")) OR AB,TI,IF("sleep duration" OR "sleep quality" OR "quality of sleep" OR "sleep measur*" OR "sleep onset" OR bedtime* OR "sleep quantity" OR "length of sleep" OR "amount of sleep" OR "sleep hour*" OR "hour of sleep" OR "sleep amount*" OR sleepiness OR sleepless*) OR AB,TI,IF("objective sleep*" OR "subjective sleep*"))) OR (((TITLE-ABS ("blood pressure" OR hypertens* OR "heart rate variation*" OR "heart rate variabl*" OR hrv OR "body mass index*" OR bmi OR "waist circumference" OR obesit* OR overweight OR obese OR cytokine* OR chemokine* OR interferon* OR "interleukin 1 receptor antagonist protein*" OR interleukin* OR lymphokine* OR monokine* OR "transforming growth factor beta" OR "tumor necrosis factor*" OR "C-Reactive Protein*" OR glucocorticoid* OR lipid* OR "fatty acid*" OR lipopolysaccharide* OR triglyceride* OR cholesterol* OR hdl OR vldl OR ldl OR "blood glucose" OR "blood sugar*" OR "gut microbiota" OR "gastrointestinal microbiome*" OR "physiologic* biomarker*" OR "biologic biomarker*" OR tnf-a OR il-6 OR a1c OR "Blood Sedimentation" OR "erythrocyte sedimentation*")) OR (TITLE-ABS ((plasma OR saliva OR hair OR serum) AND cortisol))))) |

**Database: CINAHL Plus with Full Text
Provider: EBSCOhost**

Limits: English, Years 2013-2023
Exclusions: Dissertations

|  | Concept | Search Strategy |
| --- | --- | --- |
| #1 | Health Care Professionals | (MH "Health Personnel+") OR TI ( allergist* or anesthesiologist* or anesthetist* or attending* or cardiologist* or clinician* or dermatologist* or diabetologist* or doctor* or emergency-physician* or endocrinologist* or fellow or fellows or gastroenterologist* or general-practitioner* or geriatrician* or gynecologist* or gynaecologist* OR hematologist* or haematologist* or hepatologist* or hospitalist* OR immunologist* OR internist* OR intensivist* OR internist* or medical-resident* or neonatologist* or nephrologist* or neurologist* or neurosurgeon* or nurse or nurses or obstetrician* or oncologist* or ophthalmologist* or oral-surgeon* or osteopath or osteopaths or otolaryngologist* or pediatrician* or paediatrician* or pharmacist* or phlebotomist* or physician* or podiatrist* or psychologist* or psychiatrist* or pulmonologist* or rheumatologist* or social-worker* or surgeon* or therapist* or toxicologist* or urologist* or allied-health-profession* or allied-health- personnel* or community-health-worker* or counsellor* or counselor* or emergency-medical-technician* or "first responder*" OR health-educator* or healthcare assistant* OR health-care-assistant* or healthcare-support-worker* OR health-care- support-worker* or home-health-aide* or licensed-practical- nurse* or LPN or nurse*-aide* or occupational-therapist* or paramedic* or pediatric-assistant* OR paediatric-assistant* or pharmacy-technician* or physical-therapist* or physical-therapy-assistant* or physician-assistant* or psychiatric-aide* or respiratory-therapist* or speech-pathologist* ) OR AB ( allergist* or anesthesiologist* or anesthetist* or attending* or cardiologist* or clinician* or dermatologist* or diabetologist* or doctor* or emergency-physician* or endocrinologist* or fellow or fellows or gastroenterologist* or general-practitioner* or geriatrician* or gynecologist* or gynaecologist* OR hematologist* or haematologist* or hepatologist* or hospitalist* OR immunologist* OR internist* OR intensivist* OR internist* or medical-resident* or neonatologist* or nephrologist* or neurologist* or neurosurgeon* or nurse or nurses or obstetrician* or oncologist* or ophthalmologist* or oral-surgeon* or osteopath or osteopaths or otolaryngologist* or pediatrician* or paediatrician* or pharmacist* or phlebotomist* or physician* or podiatrist* or psychologist* or psychiatrist* or pulmonologist* or rheumatologist* or social-worker* or surgeon* or therapist* or toxicologist* or urologist*or allied-health-profession* or allied-health- personnel* or community-health-worker* or counsellor* or counselor* or emergency-medical-technician* or "first responder*" OR health-educator* or healthcare-assistant* OR health-care-assistant* or healthcare-support worker* OR health-care-support-worker* or home-health-aide* or licensed-practical- nurse* or LPN or nurse*-aide* or occupational-therapist* or paramedic* or paediatric-assistant* OR pediatric-assistant* or pharmacy-technician* or physical-therapist* or physical-therapy-assistant* or physician-assistant* or psychiatric-aide* or respiratory-therapist* or speech-pathologist* ) OR TI((health or health care or healthcare OR medical) N2 (worker* or personnel* OR practitioner* or provider* or professional* or navigator* or staff*)) OR AB (health or health care or healthcare) N2 AB((worker* or personnel* OR practitioner* or provider* or professional* or navigator* or staff*)) |
| #2 | Psychological and Physiological Stress, Traumatic Stress Disorders, & Fatigue | (MH "Mental Fatigue+") OR (MH "Fatigue")  OR (MH "Stress, Psychological+") OR (MH "Stress, Occupational+") OR (MH "Burnout, Professional+")  OR (MH "Stress Disorders, Post-Traumatic")  OR (MH "Psychological Trauma+")  OR TI ptsd OR AB ptsd OR TI burnout* OR AB burnout* OR TI burn-out* OR AB burn-out* OR TI occupational-stress* OR AB occupational-stress* OR TI job-stress* OR AB job-stress* OR TI mental-stress* OR AB mental-stress* OR TI workplace-stress* OR AB workplace-stress* OR TI exhaustion OR AB exhaustion OR TI trauma* W3 disorder* OR AB trauma* W3 disorder* OR   (MH "Stress, Physiological")  OR (MH "Oxidative Stress")  OR (MH "Stress") |
| #3 | Physiological Biomarkers | (MH "Sleep+") OR (MH "Sleep Stages+")  OR TI (bedtime* OR sleep-duration or sleep-quality or "quality of sleep" or sleep-measur* or sleep-onset or bedtime* or sleep-quantity or "length of sleep" or "amount of sleep" or sleep-hour* or "hour* of sleep" or sleep-amount* or sleepiness or sleepless* or objective sleep* or subjective sleep*) OR AB (bedtime* OR sleep-duration or sleep-quality or "quality of sleep" or sleep-measur* or sleep-onset or bedtime* or sleep-quantity or "length of sleep" or "amount of sleep" or sleep-hour* or "hour* of sleep" or sleep-amount* or sleepiness or sleepless* or objective sleep* or subjective sleep*) OR (MH "Blood Pressure+") OR (MH "Hypertension") OR TI blood-pressure* OR AB blood-pressure* OR TI hypertens* OR AB hypertens* OR  (MH "Heart Rate Variability")  OR TI heart rate variation* OR AB heart rate variation* OR TI hrv OR AB hrv  (MH "Body Mass Index") OR (MH "Waist Circumference")  OR (MH "Obesity") OR TI bmi OR AB bmi OR TI body-mass-index* OR AB body-mass-index* OR TI obese OR AB obese OR TI obesit* OR AB obesit* OR TI overweight OR AB overweight OR   (MH "Cytokines") OR (MH "Chemokines+") OR (MH "Tumor Necrosis Factor") OR (MH "Transforming Growth Factor beta") OR (MH "Monokines+") OR (MH "Lymphokines+") OR (MH "Interferons")  OR TI interleukin-1-receptor-antagonist protein* OR AB interleukin-1-receptor- antagonist-protein*  OR (MH "Interleukins+")  OR MH "C-Reactive Protein")  OR (MH "Glucocorticoids")  OR (MH "Lipids") OR (MH "Fatty Acids+") OR (MH "Lipopolysaccharides")  OR (MH "Lipoproteins, HDL Cholesterol") OR (MH "Cholesterol") OR (MH "Cholesterol, Dietary") OR (MH "Lipoproteins, LDL Cholesterol")  OR (MH "Triglycerides")  OR (MH "Blood Glucose")  OR (MH "Microbiota+")  OR TI gastrointestinal-microbiome OR AB gastrointestinal-microbiome OR TI microbiome* OR AB microbiome* OR TI gut- microbiota* OR AB gut-microbiota*  OR (MH "Biological Markers+")  OR TI physiologic*-biomarker* OR AB physiologic*-biomarker* OR TI biologic*-biomarker* OR AB biologic*-biomarker* OR TI cytokine* OR AB cytokine* OR TI TNF-a OR AB TNF-a OR TI c-reactive OR AB c-reactive OR TI triglyceride* OR AB triglyceride*  OR TI il-6 OR AB il-6 OR TI interleukin-6 OR AB interleukin-6 OR TI lipid* OR AB lipid* OR TI ldl OR AB ldl OR TI hdl OR AB hdl OR TI vldl OR AB vldl  OR TI cholesterol* OR AB cholesterol* OR TI blood-glucose OR AB blood-glucose OR TI a1c OR AB a1c OR TI blood-sugar* OR AB blood-sugar*  OR (MH "Blood Sedimentation")  OR TI erythrocyte-sedimentation-rate* OR AB erythrocyte-sedimentation-rate*  OR TI ( plasma OR saliva OR hair OR serum ) AND TI cortisol  OR AB ( plasma OR saliva OR hair OR serum ) AND AB cortisol |

**Database: Embase: Excerpta Medica Database
Provider: Elsevier**

Limits: English, Years 2013-2023
Exclusions: Conference Abstracts

|  | Concept | Search Strategy |
| --- | --- | --- |
| #1 | Health Care Professionals | 'allied health profession*':ab,ti,kw OR 'allied health personnel*':ab,ti,kw OR 'community health worker*':ab,ti,kw OR counsellor*:ab,ti,kw OR counselor*:ab,ti,kw OR 'emergency medical technician*':ab,ti,kw OR 'first responder*':ab,ti,kw OR 'health educator*':ab,ti,kw OR 'healthcare assistant*':ab,ti,kw OR 'healthcare support worker*':ab,ti,kw OR 'home health aide*':ab,ti,kw OR 'licensed practical nurse*':ab,ti,kw OR lpn:ab,ti,kw OR 'nurse* aides':ab,ti,kw OR 'occupational therapist*':ab,ti,kw OR paramedic*:ab,ti,kw OR 'paediatric assistant*':ab,ti,kw OR 'pediatric assistant*':ab,ti,kw OR 'pharmacy technician*':ab,ti,kw OR 'physical therapist*':ab,ti,kw OR 'physical therapy assistant*':ab,ti,kw OR 'physician assistant*':ab,ti,kw OR 'psychiatric aide*':ab,ti,kw OR 'respiratory therapist*':ab,ti,kw OR 'speech pathologist*':ab,ti,kw  OR  'paramedical personnel'/exp OR 'nurse'/exp OR 'pharmacist'/exp OR 'radiographer'/exp  OR  allergist*:ab,ti,kw OR anesthesiologist*:ab,ti,kw OR anesthetist*:ab,ti,kw OR attending*:ab,ti,kw OR cardiologist*:ab,ti,kw OR clinician*:ab,ti,kw OR dermatologist*:ab,ti,kw OR diabetologist*:ab,ti,kw OR doctor*:ab,ti,kw OR 'emergency physician*':ab,ti,kw OR endocrinologist*:ab,ti,kw OR fellow:ab,ti,kw OR fellows:ab,ti,kw OR gastroenterologist*:ab,ti,kw OR 'general practitioner*':ab,ti,kw OR geriatrician*:ab,ti,kw OR gynecologist*:ab,ti,kw OR hematologist*:ab,ti,kw OR haematologist*:ab,ti,kw OR hospitalist*:ab,ti,kw OR immunologist*:ab,ti,kw OR intensivist*:ab,ti,kw OR internist*:ab,ti,kw OR 'medical resident*':ab,ti,kw OR neonatologist*:ab,ti,kw OR nephrologist*:ab,ti,kw OR neurologist*:ab,ti,kw OR neurosurgeon*:ab,ti,kw OR nurse:ab,ti,kw OR nurses:ab,ti,kw OR obstetrician*:ab,ti,kw OR oncologist*:ab,ti,kw OR ophthalmologist*:ab,ti,kw OR 'oral surgeon*':ab,ti,kw OR osteopath:ab,ti,kw OR osteopaths:ab,ti,kw OR otolaryngologist*:ab,ti,kw OR pediatrician*:ab,ti,kw OR pharmacist*:ab,ti,kw OR phlebotomist*:ab,ti,kw OR physician*:ab,ti,kw OR podiatrist*:ab,ti,kw OR psychologist*:ab,ti,kw OR psychiatrist*:ab,ti,kw OR pulmonologist*:ab,ti,kw OR rheumatologist*:ab,ti,kw OR 'social worker*':ab,ti,kw OR surgeon*:ab,ti,kw OR therapist*:ab,ti,kw OR toxicologist*:ab,ti,kw OR urologist*:ab,ti,kw  OR  'health care personnel'/exp OR 'advanced practice provider'/exp OR 'anesthesist'/exp OR 'health educator'/exp OR 'hospital personnel'/exp OR 'psychologist'/de OR 'social worker'/de OR 'medical specialist'/de OR 'medical staff'/de OR 'physician'/exp OR 'cardiologist'/exp OR 'gynecologist'/exp OR 'hematologist'/exp OR 'obstetrician'/exp OR 'oncologist'/exp OR 'orthopedic specialist'/exp OR 'pathologist'/exp OR 'pediatrician'/exp OR 'radiologist'/exp OR 'interventional radiologist'/exp OR 'neuroradiologist'/exp OR 'surgeon'/exp OR 'urologist'/exp  OR  (health OR 'health care' OR healthcare OR medical) NEAR/2 (personnel* OR practitioner* OR provider* OR professional* OR navigator* OR staff* OR worker*) |
| #2 | Psychological and Physiological Stress, Traumatic Stress Disorders, & Fatigue | 'mental stress'/exp OR 'burnout'/de OR 'professional burnout'/de OR 'job stress'/exp OR 'critical incident stress'/de OR 'posttraumatic stress disorder'/de OR 'chronic stress'/exp OR 'mental fatigue'/de OR 'fatigue'/de OR 'exhaustion'/de OR 'psychotrauma'/de  OR  'psychological stress*':ab,ti,kw OR 'psychological trauma':ab,ti,kw OR 'burn out*':ab,ti,kw OR burnout*:ab,ti,kw OR ptsd:ab,ti,kw OR 'occupational stress*':ab,ti,kw OR 'workplace stress*':ab,ti,kw OR (trauma* NEXT/3 disorder*)  OR  'physiological stress'/exp OR 'oxidative stress'/de |
| #3 | Physiological Biomarkers | (plasma:ab,ti,kw OR saliva:ab,ti,kw OR hair:ab,ti,kw OR serum:ab,ti,kw) AND cortisol:ab,ti,kw  OR  'physiologic* biomarker*':ab,ti,kw OR 'biologic* biomarker*':ab,ti,kw OR cytokine*:ab,ti,kw OR 'tnf a':ab,ti,kw OR 'il 6':ab,ti,kw OR 'c reactive protein*':ab,ti,kw OR lipid*:ab,ti,kw OR ldl:ab,ti,kw OR hdl:ab,ti,kw OR vdl:ab,ti,kw OR cholesterol:ab,ti,kw OR a1c:ab,ti,kw OR microbiome*:ab,ti,kw OR 'blood sugar*':ab,ti,kw OR 'gut microbiota*':ab,ti,kw  OR  'erythrocyte sedimentation rate'/de OR 'blood sedimentation*':ab,ti,kw  OR  'microflora'/de OR microbiota:ab,ti,kw OR 'intestine flora'/exp OR 'gastrointestinal microbiome':ab,ti,kw  OR  'glucose blood level'/de OR 'blood glucose':ab,ti,kw  OR  'triacylglycerol'/de OR triglyceride*:ab,ti,kw  OR  'high density lipoprotein cholesterol'/de OR 'low density lipoprotein cholesterol'/de OR 'very low density lipoprotein cholesterol'/de  OR  'c reactive protein'/de OR 'glucocorticoid'/de OR 'lipid'/de OR 'fatty acid'/exp OR 'essential fatty acid'/exp OR 'hydroxy fatty acid'/exp OR 'long chain fatty acid'/exp OR 'medium chain fatty acid'/exp OR 'short chain fatty acid'/exp OR 'unsaturated fatty acid'/exp OR 'lipopolysaccharide'/exp  OR  'lymphokine'/de OR 'leukocyte migration inhibition factor'/de OR 'lymphotoxin'/de OR 'macrophage activating factor'/de OR 'macrophage migration inhibition factor'/de OR 'suppressor factor'/de OR 'transfer factor'/de OR 'monokine'/de OR 'tumor necrosis factor'/de OR 'transforming growth factor beta'/de  OR  'interleukin 26'/de OR 'interleukin 27'/de OR 'interleukin 28'/de OR 'interleukin 28b'/de OR 'interleukin 3'/de OR 'interleukin 31'/de OR 'interleukin 32'/de OR 'interleukin 33'/de OR 'interleukin 34'/de OR 'interleukin 35'/de OR 'interleukin 37'/de OR 'interleukin 4'/de OR 'interleukin 5'/de OR 'interleukin 7'/de OR 'interleukin 9'/de  OR  'cytokine'/de OR 'chemokine'/exp OR 'interferon'/exp OR 'interleukin 1 receptor blocking agent'/de OR 'interleukin derivative'/de OR 'interleukin 1'/de OR 'interleukin 10'/de OR 'interleukin 11'/de OR 'interleukin 12'/de OR 'interleukin 12p35'/de OR 'interleukin 12p40'/de OR 'interleukin 12p70'/de OR 'interleukin 13'/de OR 'interleukin 14'/de OR 'interleukin 15'/de OR 'interleukin 16'/de OR 'interleukin 17'/de OR 'interleukin 17c'/de OR 'interleukin 17f'/de OR 'interleukin 18'/de OR 'interleukin 19'/de OR 'interleukin 1alpha'/de OR 'interleukin 1beta'/de OR 'interleukin 1beta[163-171]'/de OR 'interleukin 2'/de OR 'interleukin 2 derivative'/de OR 'interleukin 20'/de OR 'interleukin 21'/de OR 'interleukin 22'/de OR 'interleukin 23'/de OR 'interleukin 23p19'/de OR 'interleukin 24'/de OR 'interleukin 25'/de  OR  bmi:ab,ti,kw OR 'body mass index*':ab,ti,kw OR obese:ab,ti,kw OR obesit*:ab,ti,kw OR overweight:ab,ti,kw  OR  'body mass'/de OR 'waist circumference'/de OR 'obesity'/de  OR  'heart rate variabl*' OR 'heart rate variation*' OR hrv:ab,ti,kw  OR  'heart rate'/de OR 'heart rate variability'/de  OR  'blood pressure*':ab,ti,kw OR hypertens*:ab,ti,kw  OR  'blood pressure'/de OR 'hypertension'/de  OR  'sleep duration':ab,ti,kw OR 'sleep quality':ab,ti,kw OR 'quality of sleep':ab,ti,kw OR 'sleep measur*':ab,ti,kw OR 'sleep onset':ab,ti,kw OR bedtime:ab,ti,kw OR 'sleep quantity':ab,ti,kw OR 'length of sleep':ab,ti,kw OR 'amount of sleep':ab,ti,kw OR 'sleep hour':ab,ti,kw OR 'hour* of sleep':ab,ti,kw OR 'sleep amount':ab,ti,kw OR sleepiness:ab,ti,kw OR sleepless*:ab,ti,kw OR 'objective sleep*':ab,ti,kw OR 'subjective sleep*':ab,ti,kw  OR  'sleep'/exp OR 'sleep stage'/exp OR 'nonrem sleep'/exp OR 'sleep parameters'/exp |

**Database: Medline via Ovid
Provider: Wolters Kluwer**

Limits: English, Years 2013-2023

|  | Concept | Search Strategy |
| --- | --- | --- |
| #1 | Health Care Professionals | exp Health Personnel/ or Social Workers/ or (((allergist* or anesthesiologist* or anesthetist* or attending* or cardiologist* or clinician* or dermatologist* or diabetologist* or doctor* or emergency physician* or endocrinologist* or fellow or fellows or gastroenterologist* or general practitioner* or geriatrician* or gynaecologist* or gynecologist* or hematologist* or haematologist* or (health or health care or healthcare or medical)) adj2 (worker* or practitioner* or personnel* or provider* or professional or navigator* or staff*)) or hepatologist* or hospitalist* or immunologist* or intensivist* or internist* or medical resident* or neonatologist* or nephrologist* or neurologist* or neurosurgeon* or nurse or nurses or obstetrician* or oncologist* or ophthalmologist* or oral surgeon* or osteopath or osteopaths or otolaryngologist* or paediatrician* or pediatrician* or pharmacist* or phlebotomist* or physician* or podiatrist* or psychologist* or psychiatrist* or pulmonologist* or rheumatologist* or social worker* or surgeon* or therapist* or toxicologist* or urologist*).mp. OR exp Allied Health Personnel/ or (allied health profession* or allied health personnel* or community health worker* or counsellor* or counselor* or emergency medical technician* or first responder* or health educator* or healthcare assistant* or healthcare support worker* or home health aide* or licensed practical nurse* or LPN or nurse* aides or occupational therapist* or paramedic* or paediatric assistant* or pediatric assistant* or pharmacy technician* or physical therapist* or physical therapy assistant* or physician assistant* or psychiatric aide* or respiratory therapist* or speech pathologist*).mp. |
| #2 | Psychological and Physiological Stress, Traumatic Stress Disorders, & Fatigue | mental fatigue/ or exp stress, psychological/ or exp burnout, psychological/ or exp occupational stress/  OR "trauma and stressor related disorders"/ or stress disorders, traumatic/ or psychological trauma/ or stress disorders, post-traumatic/ or stress disorders, traumatic, acute/  OR (psychological trauma or ptsd or burn-out* or burnout* or occupational stress* or job stress* or workplace stress* or mental stress* or exhaustion or (trauma* adj3 disorder*)).mp. [mp=title, abstract, original title, name of substance word, subject heading word, floating sub-heading word, keyword heading word, organism supplementary concept word, protocol supplementary concept word, rare disease supplementary concept word, unique identifier, synonyms]  OR stress, physiological/ or exp oxidative stress/ |
| #3 | Physiological Biomarkers | exp sleep/ or exp sleep stages/  OR (sleep duration or sleep quality or "quality of sleep" or sleep measur* or sleep onset or bedtime* or sleep quantity or "length of sleep" or "amount of sleep" or sleep hour* or "hour* of sleep" or sleep amount* or sleepiness or sleepless* or fatigue* or objective sleep* or subjective sleep*).mp. [mp=title, abstract, original title, name of substance word, subject heading word, floating sub-heading word, keyword heading word, organism supplementary concept word, protocol supplementary concept word, rare disease supplementary concept word, unique identifier, synonyms]  OR Blood Pressure OR Hypertension/ OR (blood pressure* or hypertens*).mp. [mp=title, abstract, original title, name of substance word, subject heading word, floating sub-heading word, keyword heading word, organism supplementary concept word, protocol supplementary concept word, rare disease supplementary concept word, unique identifier, synonyms]  OR  Heart Rate/  OR  (heart rate variation* or heart rate variabl* or HRV).mp. [mp=title, abstract, original title, name of substance word, subject heading word, floating sub-heading word, keyword heading word, organism supplementary concept word, protocol supplementary concept word, rare disease supplementary concept word, unique identifier, synonyms] OR body mass index/  OR Waist Circumference/  OR  Obesity/  OR Overweight/  OR (BMI or body mass index* or obese or obesit*).mp. [mp=title, abstract, original title, name of substance word, subject heading word, floating sub-heading word, keyword heading word, organism supplementary concept word, protocol supplementary concept word, rare disease supplementary concept word, unique identifier, synonyms]  OR cytokines/ or exp chemokines/ or exp interferons/ or interleukin 1 receptor antagonist protein/ or exp interleukins/ or exp lymphokines/ or exp monokines/ or exp transforming growth factor beta/ or tumor necrosis factors/   OR C-Reactive Protein/  OR Glucocorticoids/  OR lipids/ or exp fatty acids/ or exp lipopolysaccharides/ OR Cholesterol, HDL/ or Cholesterol, VLDL/ or Cholesterol/ or Cholesterol, LDL/  OR  Triglycerides/  OR  Blood Glucose/  OR  microbiota/ or gastrointestinal microbiome/  OR (physiologic biomarker* or biologic* biomarker* or cytokine* or TNF-a or IL-6 or c-reactive-protein* or lipid* or LDL or HDL or VLDL or gut micobiota or blood sugar* or cholesterol* or triglyceride* or blood glucose* or A1C or microbiome*).mp. [mp=title, abstract, original title, name of substance word, subject heading word, floating sub-heading word, keyword heading word, organism supplementary concept word, protocol supplementary concept word, rare disease supplementary concept word, unique identifier, synonyms]  OR  ((plasma or saliva or hair or serum) and cortisol).mp. [mp=title, abstract, original title, name of substance word, subject heading word, floating sub-heading word, keyword heading word, organism supplementary concept word, protocol supplementary concept word, rare disease supplementary concept word, unique identifier, synonyms]  OR Blood Sedimentation/  OR erythrocyte sedimentation rate*.mp. [mp=title, abstract, original title, name of substance word, subject heading word, floating sub-heading word, keyword heading word, organism supplementary concept word, protocol supplementary concept word, rare disease supplementary concept word, unique identifier, synonyms] |
